# Longitudinal Measurements of Inflammatory Mediators in Patients at Risk of Sepsis in the Emergency Department

**DOI:** 10.64898/2026.03.02.26347244

**Authors:** Berta Cistero, Veronica Monforte, Marta Camprubi-Rimblas, Aina Areny-Balaguero, Elena Campana-Duel, Aida Fernandez, Antonio Casabella Pernas, Elisa Nuez Zaragoza, Iu Martin, Adriana Tomas, Irune Minarro, Marta Vila, Mireia Cuevas, Mar Sanchez, Xavier Belda, Monica Lopez Rodriguez, Tiago Teles, Maria F. Savone, Carlos Stable, Patricia Salom Merce, Carla Guijarro Viudez, Juan Tajan, Gemma Goma Fernandez, Maria L. Martinez, Lina Kramer, Rombout van Amstel, Emilio Diaz Santos, Lluis Blanch, Emilio M. Gene Tous, Lieuwe DJ. Bos, Antonio Artigas Raventos, Adrian Ceccato

## Abstract

Sepsis is a complex condition with time⍰dependent evolution, and longitudinal biomarker dynamics may improve its characterisation. We hypothesised that biomarker kinetics are associated with sepsis, the intensity of organ dysfunction, and patient survival.

This single⍰centre, prospective, observational study included adult patients presenting to the Emergency Department with suspected infection and a National Early Warning Score 2 (NEWS2) ≥3. Blood samples were collected at baseline, 4 and 24 hours. Linear mixed models assessed associations between biomarker trajectories, sepsis diagnosis, and organ dysfunction severity. Joint models evaluated the predictive value of biomarker kinetics during the first 24 hours for in⍰hospital mortality.

Of 214 screened patients, 173 were analysed and 137(79%) developed sepsis. Linear mixed models showed significant time⍰dependent decreases in IL10(β −0.016, 95%CI −0.028 to −0.004), IL1RN(β −0.014, 95%CI −0.024 to −0.004), and IL6(β −0.012, 95%CI −0.024 to 0.00). Sepsis diagnosis was associated with higher IL1RN(β 0.378, 95%CI 0.153– 0.603) and TNFRSF1A(β 0.40, 95%CI 0.21–0.58); a significant interaction between sepsis and time was observed only for IL6.

Increasing SOFA scores correlated with elevated IL10(β 0.048), IL1RN(β 0.044), CCL2(β 0.046), TNFRSF1A(β 0.050), and procalcitonin(PCT; β 2.63), all p<0.01. The SOFA–time interaction was significant only for IL6 (β −0.003, 95%CI −0.005 to −0.001). Joint survival models adjusted for age and maximum SOFA identified IL8 (HR 0.655, 95%CrI 0.582– 0.728), TNFRSF1A (HR 0.505, 95%CrI 0.419–0.682), and PCT (HR 1.004, 95%CrI 1.001– 1.008) as outcome predictors.

In conclusion, sepsis and organ dysfunction severity are associated with distinct inflammatory biomarker levels and kinetics. IL6 showed a unique time⍰dependent relationship, while TNFRSF1A, IL8 and PCT dynamics were associated with survival.

---

Sepsis is a heterogeneous condition that causes millions of deaths each year ^1^. It is defined as a dysregulated inflammatory response to an infection that leads to organ dysfunction ^2^. Sepsis has been described as a time-dependent disease, since it has been shown that early bundle measures improve prognosis ^3^. However, the time zero is unclear, and there is a lack of diagnostic tools for early detection.

In recent decades, the significant increase in general knowledge about sepsis physiopathology has enabled several hypotheses to be developed aiming to improve prognosis. However, when tested in clinical studies, these treatments have failed to demonstrate clinical benefits ^4^. Profiling immune and inflammatory patterns may be a promising approach to personalising medicine, reducing the risk of harm, and increasing the likelihood of success ^5–8^. Experimental models and clinical observations have enabled us to understand how inflammatory mediators evolve in sepsis ^9^. Critically ill patients with sepsis exhibit increases in cytokine levels, followed by a subsequent decrease ^10^. However, these observations were limited to patients admitted to the intensive care unit (ICU).

A better understanding of longitudinal cytokine/biomarker dynamics could help us to characterise sepsis more effectively ^11^. Sepsis is a rapidly evolving syndrome in which the host response and organ dysfunction change markedly over time. Whether early cytokine dynamics could be related to clinical events has not been adequately addressed in previous research. Dismissing the temporal dimension of biomarkers may lead to misclassification of patients’ true risk and underestimate their potential for guiding individualised therapeutic decisions.

Pro-inflammatory cytokines, such as tumour necrosis factor (TNF), interleukin (IL)-1B and IL6, the anti-inflammatory cytokine IL10, chemokines IL-8 and C-C motif chemokine ligand 2 (CCL2), IL-1 receptor antagonist (IL1RN) and soluble TNF receptors (sTNFRs), as well as other inflammatory mediators or acute-phase reactants such as soluble triggering receptor expressed on myeloid cells-1 (STREM-1) and procalcitonin (PCT), have been proposed as biomarkers for sepsis identification or prognosis for their physiopathology role in the development of the condition^9^.

We hypothesised that biomarker levels are associated with sepsis and the severity of organ dysfunction, resulting in different dynamics over time. Biomarker kinetics may also be useful for predicting survival.

Our aims were to analyse (1) the kinetics of inflammatory mediators over time in patients coming to the emergency department by infection during the first 24 hours, (2) to understand whether sepsis might alter the trajectory of biomarkers, (3) whether the severity of sepsis, as measured by the SOFA score, were associated with biomarker values at different times, and (4) whether dynamic measures could aid survival prediction.

## METHODS

### Study Design

This was a single-center, prospective, observational study conducted at Hospital Parc Taulí de Sabadell, in collaboration with the Institut d’Investigació i Innovació Parc Taulí (I3PT).

### Study Population

We included adult patients who presented to the Emergency Department (ED) with suspected infection.

Patients were included if they fulfilled the following criteria: age ≥18 years, suspected infection with a National Early Warning Score 2 (NEWS) score ≥3^12^, and willingness to provide written informed consent. Patients were excluded if they were pregnant or breastfeeding, receiving immunosuppressive therapy, antineoplastic chemotherapy, or chronic corticosteroids (prednisone ≥10 mg/day for ≥3 weeks), had received an organ transplant, had been included in another study within the previous 60 days, had been hospitalized for >48 hours, had a life expectancy of less than 6 months, or had any limitation of therapeutic effort.

### Study Procedures

Eligible patients were assessed by ED physicians. Upon obtaining informed consent, a 4 mL blood sample was collected at baseline (0h), 4h, and 24h. Vital signs (heart rate, respiratory rate, blood pressure, oxygen saturation, body temperature, and level of consciousness) were recorded at 0h, 4h, and 24h. Laboratory markers of inflammation and organ dysfunction, including differential white blood cell count, lactate, bilirubin, and creatinine, were measured at the same time points.

The final diagnosis of sepsis was determined according to Sepsis-3 criteria^2^: suspected infection and a ≥2 point increase in the Sequential Organ Failure Assessment (SOFA)^13^ score, as confirmed independently by two blinded researchers (BC-AC). Clinical assessment included evaluation of organ dysfunction (e.g., oliguria, altered mental status, respiratory distress), as well as sepsis scoring systems (qSOFA, NEWS), and microbiological data (e.g., blood, sputum, urine, skin, and stool cultures). Patients with a confirmed non-infectious diagnosis after discharge were excluded from the study.

Blood samples were centrifuged, and plasma was separated and stored at −80°C. At the end of the study, samples were thawed and analysed for inflammatory mediators (IL1B, IL1RN, IL6, IL7, IL8, IL10, IL17A, TNF, TNF receptor superfamily member 1A (TNFRSF1A), interferon gamma (IFNG), CCL2 and CCL7) using a multiplexed bead-based immunoassay (Invitrogen PPX-12-MXRWGUZ), and PCT (FineTests ref. EH0341) and sTREM-1 (Invitrogen ref: EHTREM1) were analysed by ELISA methods. The study was purely observational and did not involve any interventions or modifications to standard clinical care. Patients were treated following local guidelines and the survival sepsis campaign recommendations ^3,14,15^.

The primary outcome was the development of sepsis, as defined above, and in-hospital mortality

This study is reported following STROBE guidelines ^16^. The institution’s Ethical Review Board approved the study (Comité de Ética de Investigación con medicamentos del Parc Taulí 2022/3004), which was performed following the ethical standards laid down in the 1964 Declaration of Helsinki and its later amendments. Written informed consent was obtained from all patients or their legally authorised surrogates.

### Statistical Analysis

#### Descriptives

Continuous variables were summarized using means, medians, standard deviations, and interquartile ranges. Categorical variables were described as frequencies and percentages. Comparisons between groups were performed using the chi-square or Fisher’s exact test for categorical variables, and the Student’s t-test or Mann-Whitney U test for continuous variables, as appropriate.

Time-to-event data were analyzed using Kaplan-Meier curves and compared using the log-rank or Gehan-Breslow-Wilcoxon tests.

#### Missing data and data transformations

Missing data was considered as missing not at random, and was not imputed. For those measures below or above the limit of quantification, we imputed the minimum or maximum value.

Due to the non-normal distribution of cytokine data, log10+1 transformation was applied before analysis.

### Analyses of biomarker kinetics and associations to survival

Biomarker trajectories over time were analysed using linear mixed-effects models. The models included fixed effects for group (sepsis or non-sepsis), time (numeric, increasing with one unit each hour), and their interaction, and random effects as patient-specific intercepts and random slopes for time.

To assess the association between organ dysfunction severity and biomarker levels, linear mixed effects models were fit for biomarker levels (except for PCT and STREM-1), with time (numeric) and SOFA score as fixed effects, and patient-specific intercepts and time slope as random effects. We also tested for interaction between SOFA and time (p-value <0.05). The NLME package ^17^ was used to fit the mixed-effects models. Changes in SOFA score and log-transformed biomarker concentrations between baseline (0 h) and 24 hours were analysed using linear regression models.

To explore the predictive value of biomarker concentrations and their kinetics for in-hospital mortality, joint models for longitudinal and time-to-event data were used. These models combined a linear mixed-effects model for each biomarker with a relative risk model for survival, including age and maximum SOFA score as potential confounders. The*JMbayes2* package was used for joint modeling ^18^.

A Bayesian approach was used to fit the joint models, with posterior distributions approximated using three MCMC chains of 300,000 iterations each. The first 60,000 iterations were discarded as burn-in, and every 100th iteration was retained to reduce autocorrelation. Trace plots of the simulated values indicated good overlap between chains, suggesting stabilization. Convergence to the posterior distribution was further assessed using the potential scale reduction factor (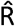), with all values close to 1, indicating convergence.

R version 4.4.0 was used for all analyses ^19^.

## RESULTS

Between September 2022 and June 2024, 214 patients were screened and accepted as participants. Of these, 196 satisfied the inclusion/exclusion criteria and 173 had data available for analysis (at least one measure of each biomarker/inflammatory mediator), as well as having their survival recorded at discharge (see Fig. 1). 137 patients (79%) developed sepsis within 24 hours of arriving at the ED and the rest were categorized as infections without sepsis n=36 (21%).

**Figure 1.**
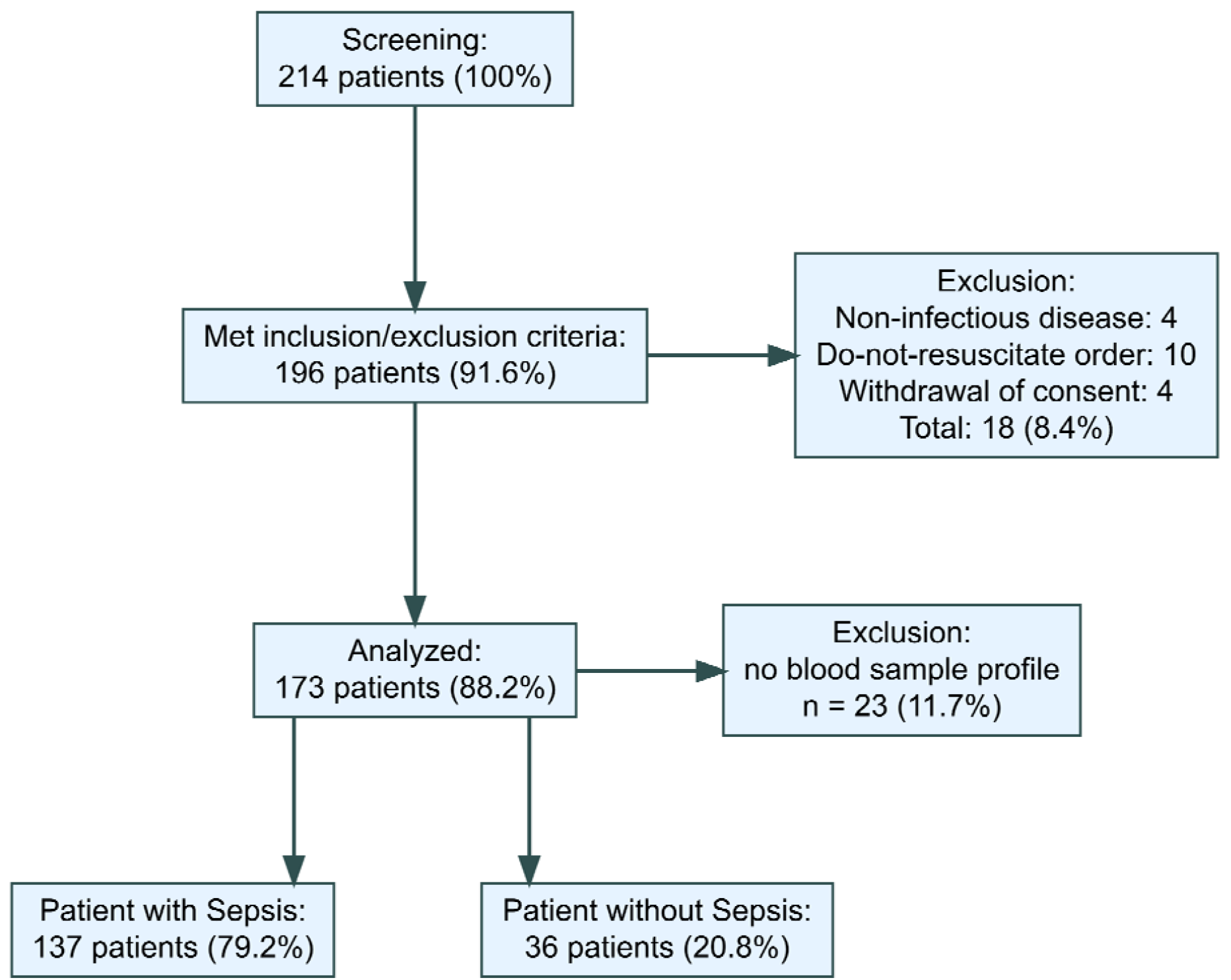
Flowchart. Study recruitment overview. Adjudication of sepsis was performed by at least two physicians based on the Sepsis-3 definition^2^.

Baseline characteristics and comorbidities are summarised in Table 1. Patients who developed sepsis were older and had similar comorbidities to those who did not. The sites of infection were mostly respiratory and urinary. The most commonly isolated pathogens (bacteremic or non-bacteremic) in non-septic patients were *E. coli* and *K. pneumoniae*, whereas in septic patients, the most commonly isolated pathogens were Enterobacteriaceae and *S. pneumoniae* (eTable 1). The outcomes for patients with sepsis were worse, requiring more ICU admissions and significantly lower survival rates with an in-hospital mortality of 29 (21%) in the group of sepsis compared with none in the non-septic group (see eFig 1).

**Table 1.** Demographic and clinical characteristics at time 0 of patients by sepsis status.

| Variable | Non-Sepsis (n=36) | Sepsis (n=137) | p-value |
| --- | --- | --- | --- |
| Age, years (median [IQR]) | 69.5 [55.75, 79] | 78 [67.00, 85] | 0.005 |
| BMI, kg/m<sup>2</sup> (median [IQR]) | 26.6 [24.2, 31.3] | 25.75 [22.52, 29.62] | 0.40 |
| Sex male, n(%) | 29 (80.6%) | 87 (63.5%) | 0.08 |
| Charlson Comorbidity Index (median [IQR]) | 5 [3, 7] | 5 [4, 7] | NS |
| Coronary disease, n(%) | 4 (11.1%) | 18 (13.1%) | 1.0 |
| Chronic Heart Failure, n(%) | 4 (11.1%) | 28 (20.4%) | 0.29 |
| Peripheral Vascular Disease, n(%) | 5 (13.9%) | 19 (13.9%) | 1.0 |
| Cerebrovascular Disease, n(%) | 5 (13.9%) | 29 (21.2%) | 0.45 |
| Chronic Pulmonary Disease, n(%) | 13 (36.1%) | 36 (26.3%) | 0.33 |
| Dementia, n(%) | 6 (16.7%) | 34 (24.8%) | 0.41 |
| Liver Disease, n(%) | 4 (11.1%) | 8 (5.8%) | 0.27 |
| Diabetes mellitus, n(%) | 8 (22.2%) | 39 (28.5%) | 0.58 |
| Chronic Kidney Disease, n(%) | 10 (27.8%) | 29 (21.2%) | 0.53 |
| Site of infection |  |  | 0.11 |
| Respiratory | 21(60%) | 55(40%) |  |
| Urinary | 8(23%) | 38(28%) |  |
| Abdominal (non-surgical) | 1(3%) | 25(18%) |  |
| Other | 5(13%) | 17(13%) |  |
| Days since the onset of symptoms, Median [Q1, Q3] | 2 [0, 3] | 2 [0, 4] | 0.99 |
| Systolic Blood Pressure mmhg (median [IQR]) | 114.5 [94.75, 130.75] | 97 [85, 114] | 0.007 |
| Diastolic Blood Pressure mmHg (median [IQR]) | 64.5 [57.25, 82.25] | 57 [49, 68] | 0.003 |
| Respiratory Rate per min (median [IQR]) | 34 [31, 36.50] | 23.50 [20, 30] | 0.021 |
| Heart Rate per min (median [IQR]) | 110 [86.5, 122.25] | 98 [78, 115.50] | 0.07 |
| SOFA Score (median [IQR]) | 1 [1, 1] | 3 [2, 5] | <0.0001 |
| APACHE Score (median [IQR]) | 9 [6.75, 11] | 12 [9.50, 16] | 0.0005 |
| NEWS2 Score (median [IQR]) | 5 [4, 6] | 6 [4, 8] | 0.06 |
| qSOFA Score (median [IQR]) | 1 [0, 1] | 1 [0.75, 1] | 0.0007 |
| SpO2/FiO2 Ratio (median [IQR]) | 442.86 [383.33, 457.14] | 419.05 [339.29, 452.38] | 0.21 |
| PaO2/FiO2 Ratio (median [IQR]) | 333.33 [264.52, 380.95] | 277.38 [196.62, 380.95] | 0.32 |
| Leukocytes 10<sup>9</sup>/L (median [IQR]) | 12.34 [9.55, 15.85] | 12.36 [7.52, 18.34] | 0.79 |
| Lymphocytes % (median [IQR]) | 8.00 [4, 12.5] | 6.00 [4, 10.25] | 0.79 |
| Neutrophils % (median [IQR]) | 85 [70.5, 89.5] | 87 [79.75, 91] | 0.21 |
| Absolute Neutrophils/<br>Lymphocytes ratio | 10.6 [7.25, 20.4] | 14.1 [7.17, 19.6] | 0.616 |
| CRP mg/dL (median [IQR]) | 6.28 [4.03, 16.12] | 11.69 [3.31, 25.84] | 0.11 |
| Lactate mmol/L (median [IQR]) | 1.68 [1.31, 2.22] | 2.51 [1.49, 3.54] | 0.005 |
| pH (median [IQR]) | 7.38 [7.34, 7.43] | 7.38 [7.33, 7.43] | 0.81 |
| paO<sub>2</sub> mmHg (median [IQR]) | 80.00 [75.00, 82.00] | 80.00 [64.38, 87.92] | 0.83 |
| paCO<sub>2</sub> mmHg (median [IQR]) | 41.00 [37.28, 44.65] | 35.20 [32.00, 43.20] | 0.0445 |
| Bicarbonate meq/L (median [IQR]) | 23.20 [22.50, 26.10] | 21.90 [19.40, 24.58] | 0.0326 |
| Creatinine mg/dL (median [IQR]) | 0.94 [0.82, 1.11] | 1.24 [0.95, 1.97] | 0.0015 |
| Glucose mg/dL (median [IQR]) | 125 [106.50, 145.25] | 143 [118, 185.50] | 0.051 |
| Bilirubin mg/dL (median [IQR]) | 0.6 [0.4, 0.75] | 0.8 [0.5, 1.35] | 0.0028 |
| Bacteremic disease | 7 (19.4%) | 35 (25.5%) | 0.0405 |
| Adequate Antibiotics, n(%) | 29 (80.6%) | 118 (86.1%) | 1.0 |
| ICU admission, n(%) | 1 (2.8%) | 29 (21.2%) | 0.019 |
| Septic Shock, n(%) | 0 (0.0%) | 19 (13.9%) | 0.014 |
| Mortality, n(%) | 0 (0.0%) | 29 (21.2%) | 0.005 |

APACHE, Acute Physiology and Chronic Health Evaluation; BMI, body mass index; CRP, C-reactive protein; FiO<sub>2</sub>, fraction of inspired oxygen; ICU, Intensive Care Unit; IQR, interquartile range; MV, mechanical ventilation; NEWS, national early warning score; PaCO<sub>2</sub>, arterial partial pressure of carbon dioxide; PaO<sub>2</sub>, partial pressure of arterial oxygen; qSOFA: quick sequential organ failure assessment score; SpO<sub>2</sub>: peripheral oxygen saturation SOFA: sequential organ failure assessment score.

A swimmer-plot summarising the description of samples obtained and follow-up is provided in eFig 2. Multiplex bead analyses for inflammatory mediators were performed in duplicate, and the quality and reading description are provided in eTable 2. The trajectories of each biomarker are shown in eFig3.

### Dynamics of biomarkers according to sepsis diagnosis

Linear mixed models were used to evaluate the associations between biomarkers over time and sepsis diagnosis. Those biomarkers for which more than 50% of the measures were below the limit of quantification were excluded, as their trend was flat (IFNG, IL1B, IL7, IL17A, CCL7, TNF) (eTable 3). In all patients with suspected infection included, IL10 (β estimate CI95% -0.016, -0.028 to -0.004), IL1RN (β estimate CI95% -0.014, -0.024 to - 0.004) and IL6 (β estimate CI95% -0.012, -0.024 to 0.00) showed a significant decrease over time (per hour). Sepsis diagnosis was associated with higher values of IL1RN (β estimate, 95% CI: 0.378, 0.153 to 0.603) and TNFRSF1A (β estimate, 95% CI: 0.40, 0.21 to 0.58). Only IL-6 showed a significant interaction between a sepsis diagnosis and time (β estimate CI95% -0.14, -0.028 to 0.00) (Fig. 2). This means that patients with sepsis experienced a steeper decrease in IL-6 levels over time (per hour). The results are summarised in eTable 3 and eFig4.

**Figure 2.**
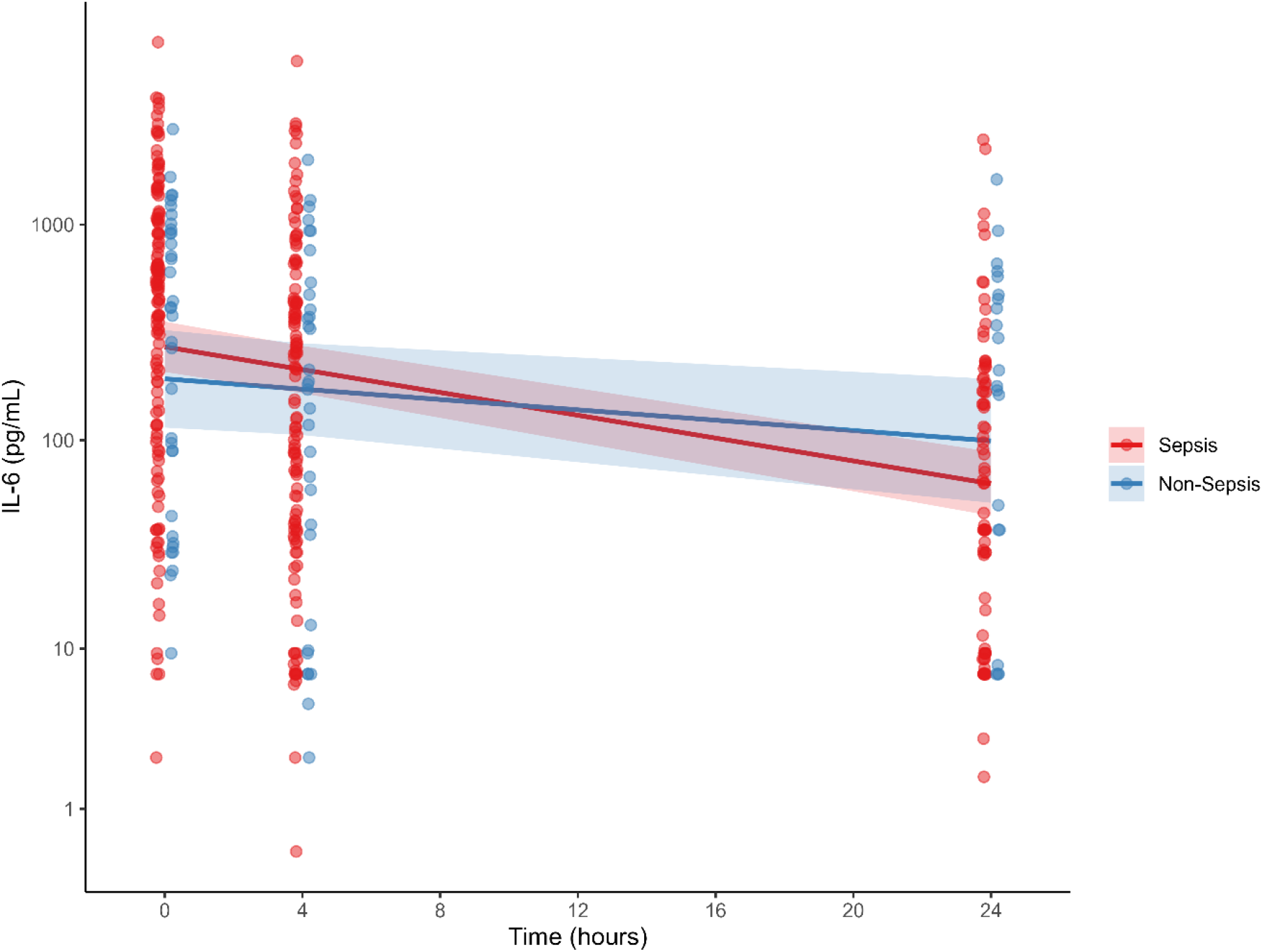
Linear mixed-effect plot for IL-6, and time according to sepsis diagnosis. Trajectories of plasma concentration IL6 during the first 24 hs by groups assigned whether or not the patients developed sepsis. Lines represent the fitted linear mixed model. Each point represents the concentration of IL-6 for each subject.

### Association of the severity of organ dysfunction on biomarkers levels

IL6 (β CI95% 0.048, 0.009 to 0.087), IL10 (β CI95% 0.048, 0.021 to 0.075), IL1RN (β CI95% 0.044, 0.017 to 0.071), CCL2(β CI95% 0.046, 0.021 to 0.071), TNFRSF1A (β CI95% 0.050, 0.030 to 0.070) and PCT (β CI95% 2.63 1.32 to 3.93) showed a significant association with organ dysfunction, where higher biomarker concentrations coincide with higher organ dysfunction. There was also a significant association between the time in hours spent in the hospital and the levels of the same biomarkers, except for PCT (eTable 3, eFig 5). Only the interaction between time and SOFA score was significant for IL6, showing that IL6 decreased with higher SOFA at 24 hours (β CI95% -0.003, -0.005 to -0.001) (Fig 3), rather than increasing as in the early stages. The results are summarised in eTable 3 and eFig. 5. A linear regression model between IL-6 delta and SOFA score delta (24-hour results minus baseline results) was performed to corroborate whether patients with a higher fall in IL6 levels have a higher fall in SOFA score. No correlation was observed between the deltas (Fig 3).

**Fig 3.**
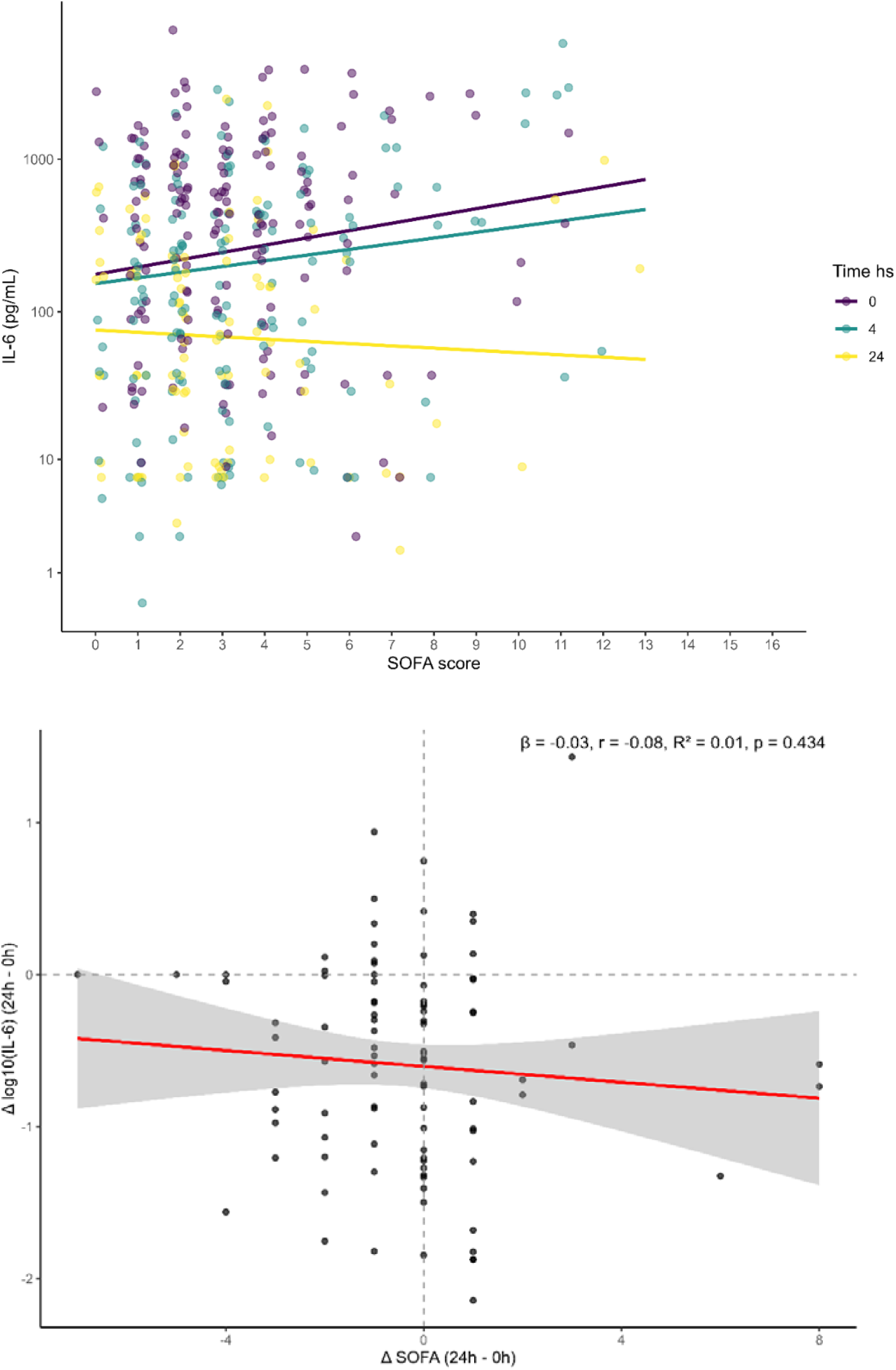
Linear mixed-effect plot for IL-6, and SOFA score according to time sampling and relation between changes of IL-6 and SOFA score at 24 hs. Above: Linear mixed model estimations of plasma concentration of IL6 according to SOFA score and time sampling. Lines represent the fitted linear mixed model. Each point represents the concentration of IL-6 for each subject. Below: Changes in SOFA score between 0 and 24 hours associated with changes in log-transformed IL-6 levels over the same period, with the regression line and corresponding slope, correlation coefficient, and coefficient of determination indicating the strength and direction of this relationship. β = regression coefficient. r= Pearson correlation coefficient. R^2^= coefficient of determination. p=p-value

### Survival prediction

Associations between the concentrations of inflammatory biomarkers and in-hospital mortality were evaluated in joint model analyses. IL8, TNFRSF1A, and PCT were associated with the hazard of death (Table 2).

**Table 2.** Results of joint modeling for longitudinal measures and time to event.

| Parameter of interest | Parameter | Hazard Ratio | Lower 95% Credible Interval | Upper 95% Credible Interval | P<sub>Bayes</sub> |
| --- | --- | --- | --- | --- | --- |
| Joint modelling evaluating IL8 | Age | 1.068 | 0.955 | 1.182 | 0.202 |
|  | Highest SOFA score | 1.415 | 1.025 | 1.856 | 0.042 |
|  | IL8 | 0.655 | 0.582 | 0.728 | 0.000 |
| Joint modelling evaluating TNFRSF1A | Age | 1.066 | 0.948 | 1.181 | 0.216 |
|  | Highest SOFA score | 1.444 | 1.042 | 1.892 | 0.035 |
|  | TNFRSF1A | 0.505 | 0.419 | 0.682 | 0.010 |
| Joint modelling evaluating PCT | Age | 1.085 | 1.032 | 1.145 | 0.002 |
|  | Highest SOFA score | 1.402 | 1.174 | 1.673 | 0.001 |
|  | PCT | 1.004 | 1.001 | 1.008 | 0.000 |

Results are obtained by joint models analyses, corrected for age, and the highest SOFA at the day of sampling. Joint model analysis was used to evaluate the longitudinal changes in plasma biomarker concentrations and the association between changes in biomarker concentration and outcomes. Joint modelling analyses two types of data: repeated measurements over time and time-to-event data (survival data). P<sub>Bayes</sub> indicates the posterior probability that the parameter differs from zero, as estimated from the joint Bayesian models. Credible intervals represent the ranges containing the true parameter value with 95% posterior probability.

Results of joint modelling, evaluating biomarkers (IL10, IL1RN, IL6, CCL2, STREM-1) that were not associated with survival probability are shown in etable4.

The joint models were tested for convergence, discrimination and calibration. The discriminatory capacity was evaluated using receiver operating characteristic (ROC) curves, which were constructed to assess the ability to predict the event at 1024 hours based on biomarker measured at 24 hours. The area under the curve for the ROC curves for each model was as follows: 0.56 for evaluating the value of log TNFRSF1A, 0.63 for evaluating the value of log IL8, and 0.60 for evaluating PCT. The Brier scores were 0.13, 0.12 and 0.16, respectively. The ROC curves and calibration plots are shown in eFigs 6-7.

## DISCUSSION

In this study, we analysed the dynamics of inflammatory markers in patients suspected of having an infection within the first 24 hours of receiving ED care. The study was developed with the aim of understanding how several factors inherent to sepsis, severity of organ dysfunction and/or time, are associated with changes in the measures of inflammatory mediators and their implications for prognosis. Our main finding comprises: IL10, IL1RN, and IL6 decreased in the first 24 hours in all included patients, while IL1RN increased with the diagnosis of sepsis. Only IL6 showed a significant change in trend when patients had a sepsis diagnosis; IL6 had a fast step decrease in those patients (Fig 2). For the association between the SOFA score and the inflammatory mediators over time, IL10, IL1RN, CCL2, TNFRSF1A, and PCT increased along with the SOFA score. Time measured in hours was associated with decreased values, with the exception of PCT, meaning that the biomarker value may be associated with a lower SOFA score on arrival or a higher SOFA score after 24 hours. In the case of IL6, the interaction between time and SOFA was significant, confirming the findings of a sepsis diagnosis, meaning that the association between SOFA and IL6 was reversed after 24 hours (Fig. 3). The ability to predict is significantly impacted by this issue. It is crucial to recognise that the harm caused by inflammation appears to be associated with the initial hours of the inflammatory response. Regarding outcome prediction, time-dependent concentrations of TNFRSF1A, IL8 and PCT during the first 24 hours were associated with the risk of in-hospital mortality and could be useful for prognosis, but with low discriminatory capacity.

The importance of our findings lies in the fact that, despite advances in the pathophysiology of sepsis, few clinical studies have focused on the kinetics of inflammatory mediators during the earliest hours of hospital care. Longitudinal analysis of data enables us to understand the behaviour of inflammatory responses and to characterise patients dynamically. Van Deuren described the kinetics of TNF, TNFRSF1A, IL1B, and IL1RN. He based his observations mainly on those in critical care patients. While IL1B and TNF showed a decrease within the first 8 hours, TNFRSF1A and IL1RN exhibited greater variability, demonstrating a smaller decrease within the first 8 hours ^10^. In another study describing the evolution of meningococcal disease, TNF and IL1B were below the limit of quantification, as in our study, and although IL6 levels increased, ex vivo production was suppressed ^20^. A study describing the kinetics of IL6 after septic shock onset found that the IL6 peak occurred near the shock onset and decreased within 24 hours ^21^. The kinetics of IL6 showed that a rapid decline within 24–48 hours was associated with successful antimicrobial therapy^22^. In our study, patients with sepsis or worst SOFA score had a significantly higher probability of decreasing IL6 levels during the first 24hs. PCT is one of the widely studied biomarkers in sepsis and has been shown to be associated with prognosis ^23–26^ and the capacity to guide antibiotic stewardship ^27–29^. PCT may increase in the first 24 hs^30^, followed by a decrease in the next days. Patients who didn’
st achieve a decrease in the levels of more than 80% have a lower survival probability ^30^. In concordance, we observed that the time-dependent values of PCT were associated with the probability of survival.

Kellum et al^31^, describing the kinetics of IL10, TNF and IL6 in a cohort of patients with pneumonia, during the first days of admission. They observed, as in our study, that there is an overlap in biomarker levels between those who developed or did not develop sepsis (former severe sepsis) (Fig 2 and eFig3), and when categorising the continuous variables, mortality was higher in those with high levels of inflammatory mediators. We observed that patients with a confirmed infection and a clear systemic response (NEWS2 >3), but not sepsis, may exhibit a similar dysfunctional immune response in terms of biomarkers as those with sepsis. This emphasises the need for a clearer definition of dysregulated responses and the concept of pre-sepsis^32^. Surprisingly, the results obtained for IL8 and TNFRSF1A predict survival with an HR below one, indicating that those with higher values in repeated measures show a better prognosis. This is opposed to what has been observed in other studies with single measures and should be confirmed.

In recent years, several endotypes or subphenotypes have been identified in sepsis and other critical illnesses, depending on the patient’s inflammatory status or genomic expression ^33–35^. Nevertheless, inflammation otherwise to some chronic conditions is not static in sepsis, changing faster even in a short time. While some retrospective analyses of clinical trial data have demonstrated the potential for improved management based on different subphenotypes or profiles (17, 18), the heterogeneity of patients and the observation that subphenotypes can change within the first few hours or days underscore the necessity for a more comprehensive understanding of the temporal evolution of inflammatory mediators ^36–39^.

Our study has strengths and limitations that should be acknowledged. The main strength lies in our protocol, which includes multiple samples within the first 24 hours, a sepsis diagnosis verified by blinded researchers, and statistical methods incorporating Linear mixed models for repeated measures ^40^ and joint models for longitudinal and time-to-event data, where we account for informative missingness of biomarker measures due to death. These methods are the most appropriate approach for assessing the impact of potential biomarkers and for dynamically updating patient prognosis ^41^. We chose three times, considering the baseline, the response to early bundles and the response to 24hs. In terms of limitations, firstly, as this is a single-centre study, the results could be influenced by the population studied. Secondly, the analysis of several inflammatory mediator measures was limited to measures above the limit of quantification. Additionally, we must consider the short time between the first two measures, which means that the sensitivity of the test could influence differences in levels. Also, not all patients are observed over the entire 24h period. Finally, the patients included in this study may have had less severe disease than those in other studies. Quarterly, we assumed time zero to be at the point of ER arrival, but this is an arbitrary decision based on the most effective way to analyse our data.

In conclusion, our comprehensive study has improved our understanding of how sepsis and organ dysfunction affect inflammatory mediators/markers during the first 24 hours after hospital admission for infection. We found that inflammation is dynamic and that the development of sepsis/organ dysfunction over time may affect inflammatory mediators and/or biomarkers within the first 24 hours of arrival at the ED in patients at risk of sepsis. IL-6 exhibits different kinetics depending on whether or not the patient develops sepsis within the first 24 hours. IL6, IL10, IL-1RN, CCL2, TNFRSF1A and PCT values were associated with the SOFA score, with different values observed at each time point (except for PCT). Finally, we demonstrated the predictive capacity of longitudinal data for survival probability. TNFRSF1A, IL8, and PCT were associated with survival and could be useful in developing prognostic tools; however, their discriminatory capacity was low.

## Data Availability

The datasets used and/or analysed during the current study are available from the corresponding author upon reasonable request.

## Abbreviations

AUCROC (AUC): Area Under the ROC Curve
CCL2: C-C motif chemokine ligand 2
CCL7: C-C motif chemokine ligand 7
CI95%: 95% Confidence Interval
ED: Emergency Department
ELISA: Enzyme-Linked Immunosorbent Assay
HR: Hazard Ratio
ICU: Intensive Care Unit
IFNG: Interferon gamma
IL1B: Interleukin 1 beta
IL1RN: Interleukin 1 receptor antagonist
IL6: Interleukin 6
IL7: Interleukin 7
IL8: Interleukin 8
IL10: Interleukin 10
IL17A: Interleukin 17A
MCMC: Markov Chain Monte Carlo
NEWS: National Early Warning Score
PCT: Procalcitonin
qSOFA: quick Sequential Organ Failure Assessment
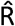 (R-hat): Potential Scale Reduction Factor
ROC: Receiver Operating Characteristic (curve)
SOFA: Sequential Organ Failure Assessment
STREM-1: Soluble Triggering Receptor Expressed on Myeloid cells-1
STROBE: Strengthening the Reporting of Observational Studies in Epidemiology
TNF: Tumor Necrosis Factor
TNFRSF1A: Tumor Necrosis Factor Receptor Superfamily Member 1A

## Ethical statement

The institution’s Ethical Review Board approved the study (Comité de Ética de Investigación con medicamentos del Parc Taulí 2022/3004), which was conducted in accordance with the ethical standards set out in the 1964 Declaration of Helsinki and its subsequent amendments. Written informed consent was obtained from all patients or their legally authorised surrogates.

## Data Availability

The datasets used and/or analysed during the current study are available from CORA.Repositori de Dades de Recerca, https://doi.org/10.34810/data3142.

## Authors’ contribution

AC & AA Study supervision. All authors reviewed and approved the manuscript for submission. **Authors’ contributions:** BC, VM, MCR, ED, EG, LB, AA and AC participated in protocol development, study design, study management, statistical analysis, and data interpretation. AF, LK, RvA, LB and AC participated in statistical analysis. BC, ACa; EN; PS; CG; JT; GG; and MLM participated in data collection. VM, MCR, AAB, and ECD participated in sample analyses. All authors read and approved the final manuscript.

## Acknowledgments

We are indebted to all our medical and nursing colleagues for their assistance and cooperation in this study.

