## Supplementary for "Longitudinal Measurements of Inflammatory Mediators in Patients at Risk of Sepsis in the Emergency Department" (from another version of this preprint: version 1, 2026-03-03)

**eFig 1. Kaplan-Meier curve, representing the survival probability according to sepsis diagnosis at the first 24 hs.**

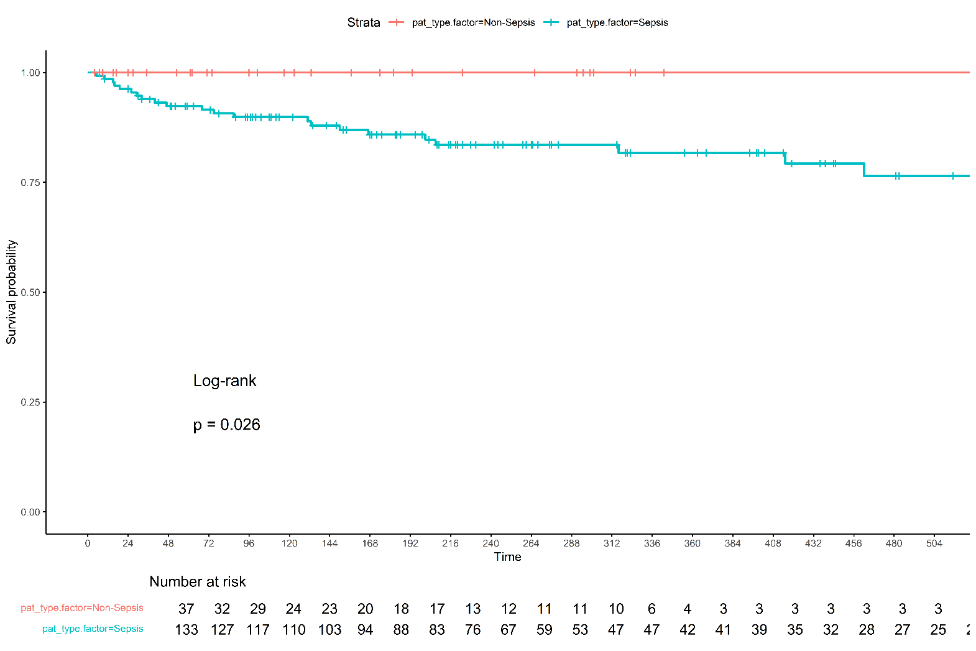

**eFig 2. Swimmer plot representing follow-up and plasma sample availability. Each line represents one subject in the study, from admission to the ED until hospital discharge or death. The color of the line indicates the patients’ status according to sepsis diagnosis.**

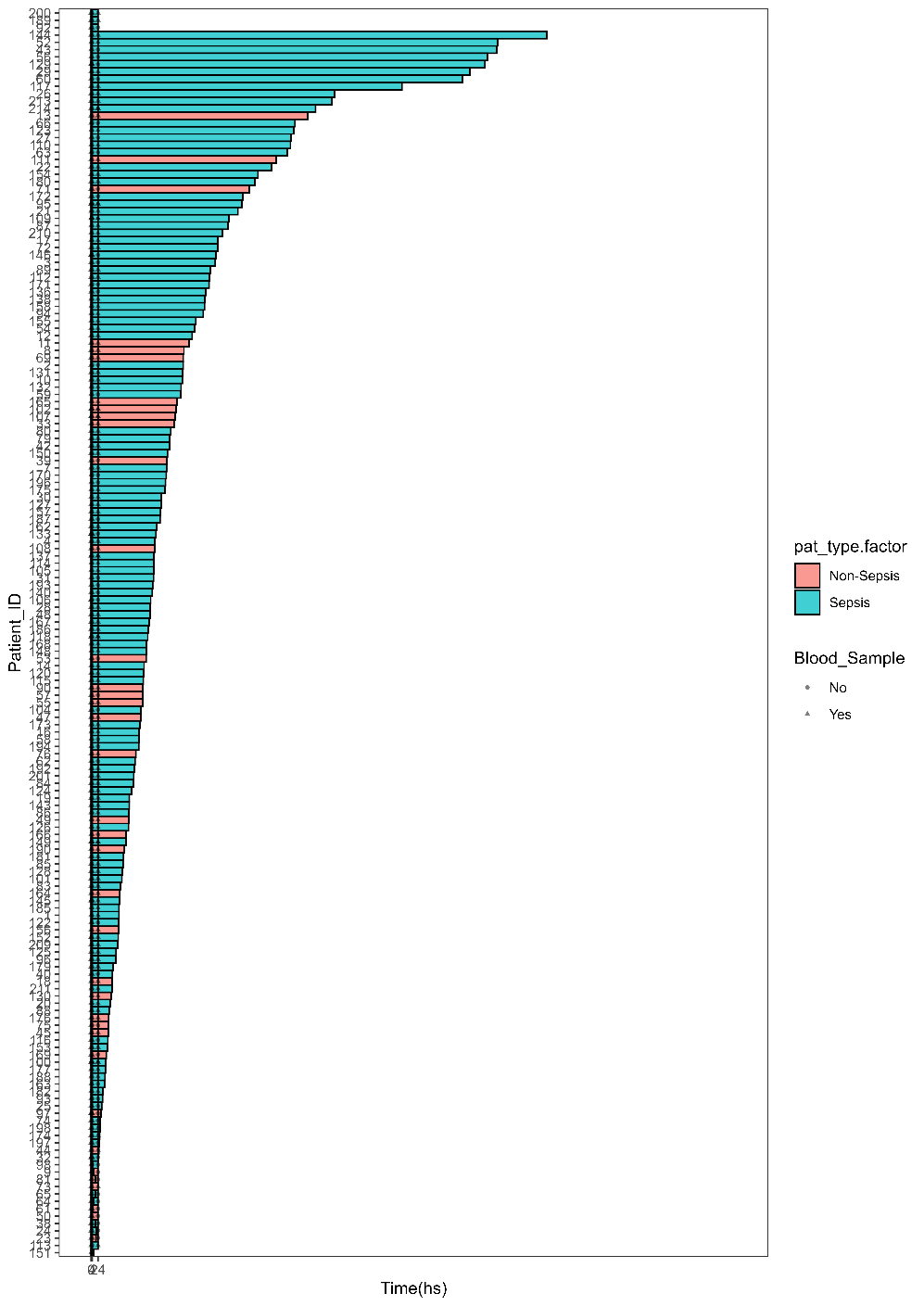

**eTable 1. Microbiologic diagnosis according to groups.**

| Microorganism isolated | Non-Sepsis n=12 | Sepsis n=58 |
| --- | --- | --- |
| *Enterococcus spp* | 1 (8.3%) | 4 (6.9%) |
| *Escherichia coli* | 5 (41.7%) | 24 (41.4%) |
| *Klebsiella pneumoniae* | 4 (33.3%) | 4 (6.9%) |
| *Legionella pneumophila* | 1 (8.3%) | 0 (0%) |
| *Neisseria meningitidis* | 1 (8.3%) | 1 (1.7%) |
| *Clostridium difficile* | 0 (0%) | 2 (3.4%) |
| *Enterobacter species* | 0 (0%) | 1 (1.7%) |
| *Haemophilus influenzae* | 0 (0%) | 1 (1.7%) |
| *Pseudomonas aeruginosa* | 0 (0%) | 2 (3.4%) |
| *Salmonella spp* | 0 (0%) | 2 (3.4%) |
| *Staphylococci spp* | 0 (0%) | 2 (3.4%) |
| *Staphylococcus aureus* | 0 (0%) | 1 (1.7%) |
| *Streptococcus pneumoniae* | 0 (0%) | 6 (10.3%) |
| *Streptococcus pyogenes* | 0 (0%) | 3 (5.2%) |
| *Other* | 0 (0%) | 5 (8.6%) |

**eTable 2. Quality of plasma Samples multiplex analysis**

|  |  | **Bead Count** | | | **Quality Check** | | | | |
| --- | --- | --- | --- | --- | --- | --- | --- | --- | --- |
| Variable | Total Samples | Good Bead Count | Low Bead Count | Low Acceptable Bead Count | Above ULQ - extrapolated based on standard curve | Above ULQ - set to non-calculated ULQ | Good quality | Under LLQ - extrapolated based on standard curve | Under LLQ - set to non-calculated LLQ |
| IFNG | 960 | 926 (96%) | 16 (1.7%) | 18 (1.9%) | 0 (0%) | 1 (0.1%) | 66 (6.9%) | 178 (19%) | 714 (74%) |
| IL10 | 960 | 910 (95%) | 20 (2.1%) | 30 (3.1%) | 0 (0%) | 0 (0%) | 611 (64%) | 231 (24%) | 118 (12%) |
| IL17A | 1,118 | 1024 (92%) | 36 (3.2%) | 58 (5.2%) | 2 (0.18%) | 0 (0%) | 204 (18%) | 185 (17%) | 727 (65%) |
| IL1B | 1,038 | 977 (94%) | 22 (2.1%) | 39 (3.8%) | 1 (0.096%) | 0 (0%) | 256 (25%) | 251 (24%) | 530 (51%) |
| IL1RN | 960 | 920 (96%) | 17 (1.8%) | 23 (2.4%) | 22 (2.3%) | 7 (0.73%) | 885 (92%) | 8 (0.83%) | 38 (4%) |
| IL6 | 960 | 879 (92%) | 30 (3.1%) | 51 (5.3%) | 0 (0%) | 2 (0.21%) | 783 (82%) | 42 (4.4%) | 133 (14%) |
| IL7 | 1,196 | 1090 (91%) | 64 (5.4%) | 42 (3.5%) | 1 (0.084%) | 0 (0%) | 563 (47%) | 406 (34%) | 226 (19%) |
| IL8 | 1,038 | 972 (94%) | 26 (2.5%) | 40 (3.9%) | 2 (0.19%) | 0 (0%) | 1008 (97%) | 25 (2.4%) | 3 (0.29%) |
| CCL2 | 960 | 878 (91%) | 38 (4%) | 44 (4.6%) | 0 (0%) | 0 (0%) | 912 (95%) | 23 (2.4%) | 25 (2.6%) |
| MCP-3 | 1,198 | 1157 (97%) | 17 (1.4%) | 24 (2%) | 0 (0%) | 0 (0%) | 380 (32%) | 62 (5.2%) | 754 (63%) |
| TNF-A | 1,038 | 1006 (97%) | 16 (1.5%) | 16 (1.5%) | 0 (0%) | 0 (0%) | 430 (41%) | 449 (43%) | 159 (15%) |
| TNFRSF1A | 1,200 | 1099 (92%) | 62 (5.2%) | 39 (3.2%) | 4 (0.33%) | 8 (0.67%) | 1157 (97%) | 26 (2.2%) | 3 (0.25%) |

Abbreviations: IL, interleukin; TNF-A, tumor necrosis factor alpha; TNFRSF1A, tumor necrosis factor receptor 1; IFNG, interferon gamma; MCP, monocyte chemoattractant protein;

**eFig 3. Trajectories for inflammatory mediators.**

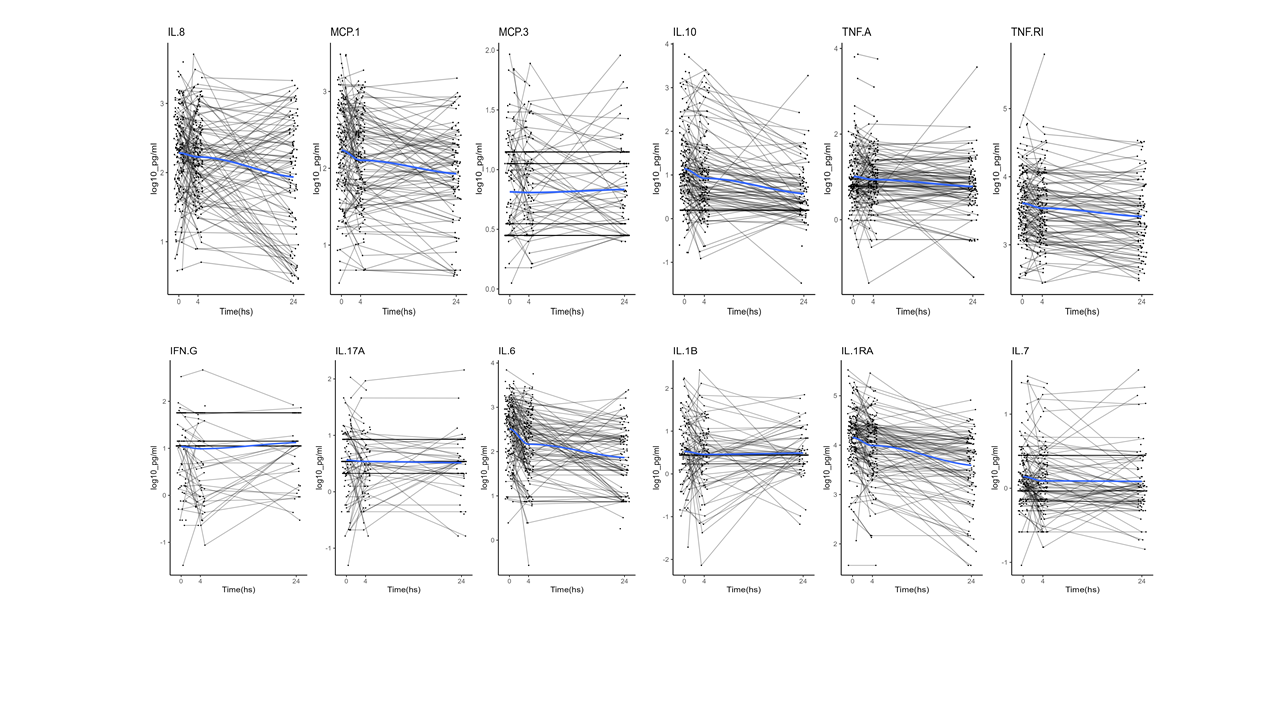

**eFig4. Linear mixed-effect plots, concentration of inflammatory mediators, and p-values of the interaction between sepsis diagnosis, concentration, and Time.**

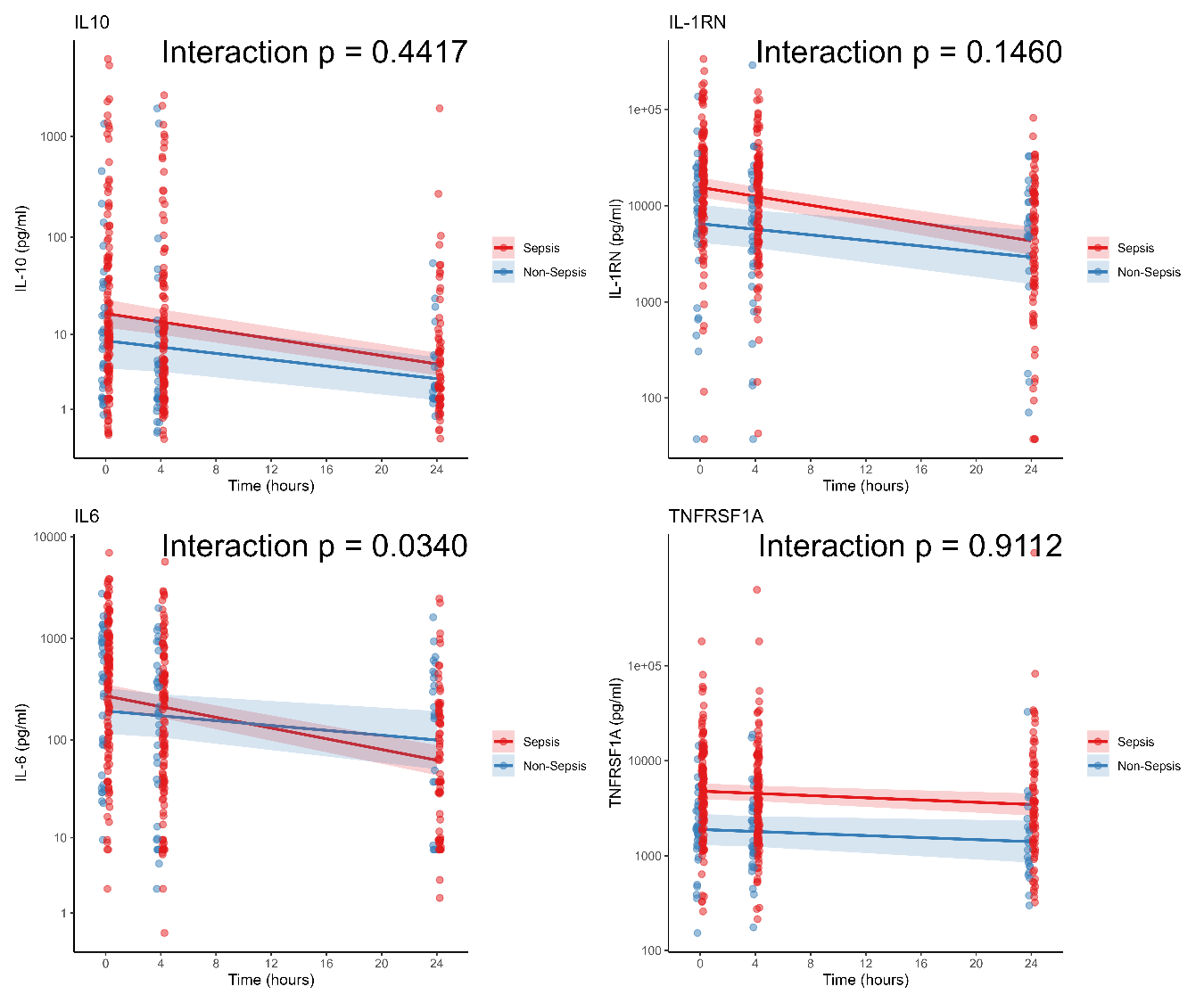

Trajectories of plasma concentration for IL10, IL-1RN, IL6, and TNFRSF1A during the first 24 hs by groups assigned whether or not the patients developed sepsis. Lines represent the fitted linear mixed model. Each point represents the concentration of IL-6 for each subject.

**eFig 5. Linear mixed-effect plots, with the predicted concentrations of each biomarker according to SOFA score and Time point.**
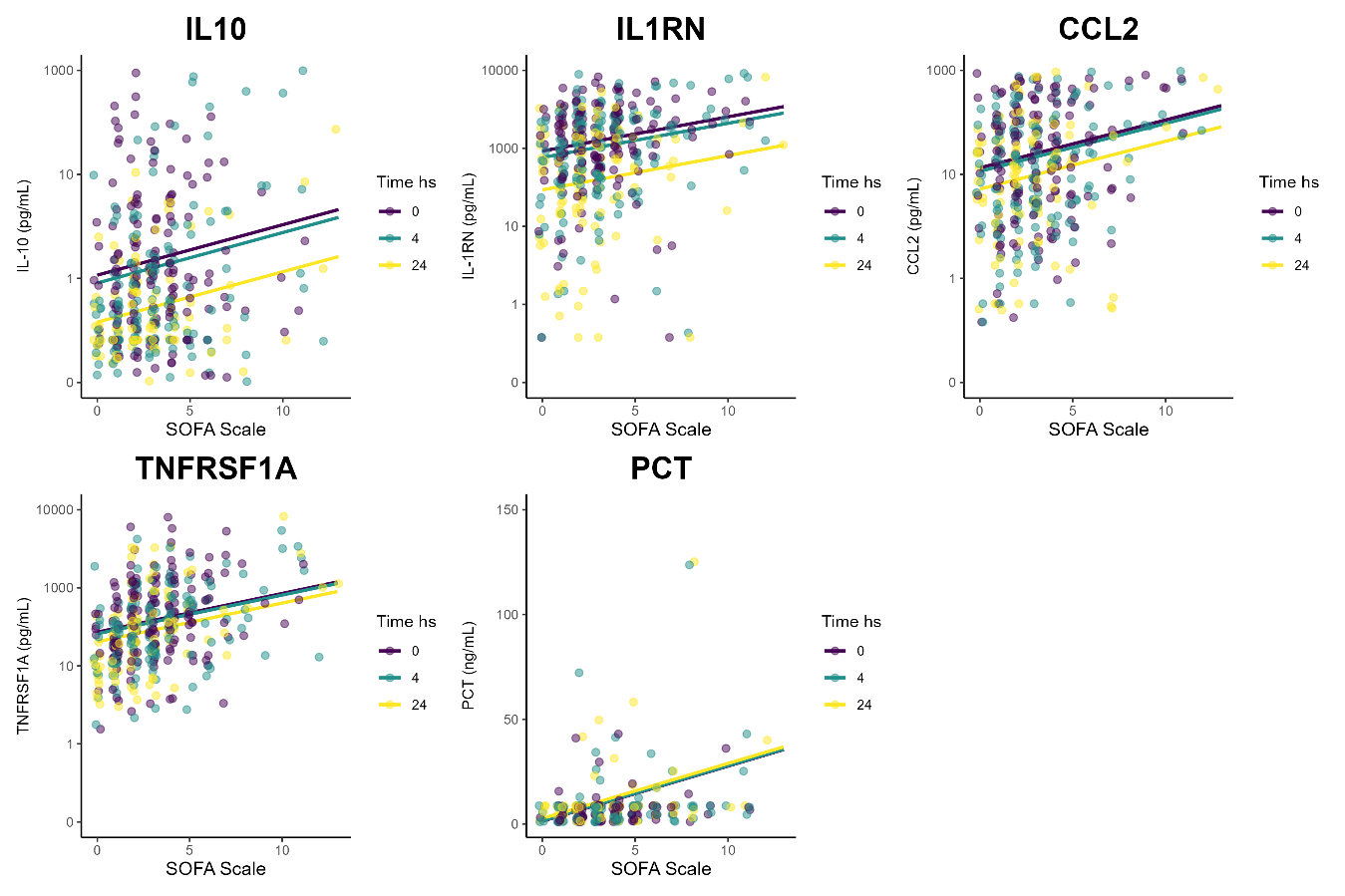

Linear mixed model estimations of plasma concentration of IL10, IL1RN, CCL2, TNFRSF1A, and PCT according to SOFA score and time sampling. Lines represent the fitted linear mixed model. Each point represents the concentration of IL-6 for each subject.

eTable 3**. Fixed and random effects estimates from mixed-effects models.**

| **Biomarker** | **Effect** | **Term** | **Beta estimate** | **2.5% confidence limit** | **97.5% confidence limit** | **p value** |
| --- | --- | --- | --- | --- | --- | --- |
| IFNG | Fixed | (Intercept) | 1.132 | 0.981 | 1.283 | <2e-16 |
| IFNG | Fixed | Sepsis diagnosis | -0.067 | -0.236 | 0.102 | 0.443 |
| IFNG | Fixed | Time | 0.003 | -0.001 | 0.007 | 0.188 |
| IFNG | Fixed | Time:Sepsis diagnosis | -0.003 | -0.009 | 0.003 | 0.245 |
| **IFNG** | **Random** | **(Intercept)** | **0.434** |  |  |  |
| **IFNG** | **Random** | **Residual** | **0.186** |  |  |  |
| **IFNG** | **Random** | **Time** | **0.005** |  |  |  |
| IL10 | Fixed | (Intercept) | 0.980 | 0.706 | 1.254 | 2.46e-11 |
| IL10 | Fixed | Sepsis diagnosis | 0.269 | -0.039 | 0.577 | 0.08897 |
| IL10 | Fixed | Time | -0.016 | -0.028 | -0.004 | 0.00876 |
| IL10 | Fixed | Time:Sepsis diagnosis | -0.005 | -0.019 | 0.009 | 0.44173 |
| **IL10** | **Random** | **(Intercept)** | **0.792** |  |  |  |
| **IL10** | **Random** | **Residual** | **0.331** |  |  |  |
| **IL10** | **Random** | **Time** | **0.025** |  |  |  |
| IL17A | Fixed | (Intercept) | 0.641 | 0.539 | 0.743 | <2e-16 |
| IL17A | Fixed | Sepsis diagnosis | 0.049 | -0.065 | 0.163 | 0.399 |
| IL17A | Fixed | Time | -0.002 | -0.006 | 0.002 | 0.527 |
| IL17A | Fixed | Time:Sepsis diagnosis | 0.001 | -0.005 | 0.007 | 0.778 |
| **IL17A** | **Random** | **(Intercept)** | **0.269** |  |  |  |
| **IL17A** | **Random** | **Residual** | **0.164** |  |  |  |
| **IL17A** | **Random** | **Time** | **0.006** |  |  |  |
| IL1B | Fixed | (Intercept) | 0.743 | 0.614 | 0.872 | <2e-16 |
| IL1B | Fixed | Sepsis diagnosis | -0.085 | -0.232 | 0.062 | 0.254 |
| IL1B | Fixed | Time | -0.002 | -0.008 | 0.004 | 0.588 |
| IL1B | Fixed | Time:Sepsis diagnosis | 0.000 | -0.008 | 0.008 | 0.919 |
| **IL1B** | **Random** | **(Intercept)** | **0.333** |  |  |  |
| **IL1B** | **Random** | **Residual** | **0.264** |  |  |  |
| **IL1B** | **Random** | **Time** | **0.008** |  |  |  |
| IL1RN | Fixed | (Intercept) | 3.812 | 3.612 | 4.012 | < 2e-16 |
| IL1RN | Fixed | Sepsis diagnosis | 0.378 | 0.153 | 0.603 | 0.00120 |
| IL1RN | Fixed | Time | -0.014 | -0.024 | -0.004 | 0.00696 |
| IL1RN | Fixed | Time:Sepsis diagnosis | -0.009 | -0.021 | 0.003 | 0.14601 |
| **IL1RN** | **Random** | **(Intercept)** | **0.563** |  |  |  |
| **IL1RN** | **Random** | **Residual** | **0.285** |  |  |  |
| **IL1RN** | **Random** | **Time** | **0.020** |  |  |  |
| IL6 | Fixed | (Intercept) | 2.288 | 2.061 | 2.515 | <2e-16 |
| IL6 | Fixed | Sepsis diagnosis | 0.148 | -0.107 | 0.403 | 0.2562 |
| IL6 | Fixed | Time | -0.012 | -0.024 | 0.000 | 0.0483 |
| IL6 | Fixed | Time:Sepsis diagnosis | -0.014 | -0.028 | 0.000 | 0.0340 |
| **IL6** | **Random** | **(Intercept)** | **0.591** |  |  |  |
| **IL6** | **Random** | **Residual** | **0.442** |  |  |  |
| **IL6** | **Random** | **Time** | **0.014** |  |  |  |
| IL7 | Fixed | (Intercept) | 0.356 | 0.278 | 0.434 | <2e-16 |
| IL7 | Fixed | Sepsis diagnosis | 0.060 | -0.028 | 0.148 | 0.183 |
| IL7 | Fixed | Time | -0.002 | -0.006 | 0.002 | 0.401 |
| IL7 | Fixed | Time:Sepsis diagnosis | 0.000 | -0.004 | 0.004 | 0.956 |
| **IL7** | **Random** | **(Intercept)** | **0.215** |  |  |  |
| **IL7** | **Random** | **Residual** | **0.118** |  |  |  |
| **IL7** | **Random** | **Time** | **0.007** |  |  |  |
| IL.8 | Fixed | (Intercept) | 2.162 | 1.980 | 2.344 | <2e-16 |
| IL.8 | Fixed | Sepsis diagnosis | 0.155 | -0.051 | 0.361 | 0.140 |
| IL.8 | Fixed | Time | -0.004 | -0.018 | 0.010 | 0.581 |
| IL.8 | Fixed | Time:Sepsis diagnosis | -0.011 | -0.025 | 0.003 | 0.134 |
| **IL.8** | **Random** | **(Intercept)** | **0.443** |  |  |  |
| **IL.8** | **Random** | **Residual** | **0.413** |  |  |  |
| **IL.8** | **Random** | **Time** | **0.020** |  |  |  |
| CCL2 | Fixed | (Intercept) | 2.045 | 1.849 | 2.241 | <2e-16 |
| CCL2 | Fixed | Sepsis diagnosis | 0.216 | -0.004 | 0.436 | 0.0557 |
| CCL2 | Fixed | Time | -0.008 | -0.018 | 0.002 | 0.0734 |
| CCL2 | Fixed | Time:Sepsis diagnosis | -0.001 | -0.011 | 0.009 | 0.8572 |
| **CCL2** | **Random** | **(Intercept)** | **0.551** |  |  |  |
| **CCL2** | **Random** | **Residual** | **0.276** |  |  |  |
| **CCL2** | **Random** | **Time** | **0.017** |  |  |  |
| CCL7 | Fixed | (Intercept) | 0.853 | 0.747 | 0.959 | <2e-16 |
| CCL7 | Fixed | Sepsis diagnosis | 0.030 | -0.090 | 0.150 | 0.618 |
| CCL7 | Fixed | Time | -0.002 | -0.006 | 0.002 | 0.407 |
| CCL7 | Fixed | Time:Sepsis diagnosis | 0.002 | -0.002 | 0.006 | 0.531 |
| **CCL7** | **Random** | **(Intercept)** | **0.298** |  |  |  |
| **CCL7** | **Random** | **Residual** | **0.148** |  |  |  |
| **CCL7** | **Random** | **Time** | **0.006** |  |  |  |
| TNF.A | Fixed | (Intercept) | 1.042 | 0.852 | 1.232 | <2e-16 |
| TNF.A | Fixed | Sepsis diagnosis | 0.023 | -0.191 | 0.237 | 0.831 |
| TNF.A | Fixed | Time | -0.005 | -0.013 | 0.003 | 0.278 |
| TNF.A | Fixed | Time:Sepsis diagnosis | -0.001 | -0.011 | 0.009 | 0.800 |
| **TNF.A** | **Random** | **(Intercept)** | **0.540** |  |  |  |
| **TNF.A** | **Random** | **Residual** | **0.262** |  |  |  |
| **TNF.A** | **Random** | **Time** | **0.016** |  |  |  |
| TNFRSF1A | Fixed | (Intercept) | 3.277 | 3.112 | 3.442 | < 2e-16 |
| TNFRSF1A | Fixed | Sepsis diagnosis | 0.403 | 0.219 | 0.587 | 3.12e-05 |
| TNFRSF1A | Fixed | Time | -0.005 | -0.013 | 0.003 | 0.183 |
| TNFRSF1A | Fixed | Time:Sepsis diagnosis | -0.001 | -0.011 | 0.009 | 0.911 |
| **TNFRSF1A** | **Random** | **(Intercept)** | **0.475** |  |  |  |
| **TNFRSF1A** | **Random** | **Residual** | **0.176** |  |  |  |
| **TNFRSF1A** | **Random** | **Time** | **0.016** |  |  |  |
| PCT | Fixed | (Intercept) | 2.064 | -9.261 | 13.389 | 0.721 |
| PCT | Fixed | Sepsis diagnosis | 10.252 | -2.482 | 22.986 | 0.116 |
| PCT | Fixed | Time | 0.025 | -0.463 | 0.513 | 0.920 |
| PCT | Fixed | Time:Sepsis diagnosis | -0.023 | -0.568 | 0.522 | 0.934 |
| **PCT** | **Random** | **(Intercept)** | **31.537** |  |  |  |
| **PCT** | **Random** | **Residual** | **16.839** |  |  |  |
| **PCT** | **Random** | **Time** | **0.816** |  |  |  |
| STREM-1 | Fixed | (Intercept) | 819.918 | 206.752 | 1,433.084 | 0.00942 |
| STREM-1 | Fixed | Sepsis diagnosis | -645.281 | -1,324.991 | 34.429 | 0.06484 |
| STREM-1 | Fixed | Time | 2.447 | -0.589 | 5.483 | 0.11580 |
| STREM-1 | Fixed | Time:Sepsis diagnosis | -1.730 | -5.086 | 1.626 | 0.31351 |
| **STREM-1** | **Random** | **(Intercept)** | **1,615.321** |  |  |  |
| **STREM-1** | **Random** | **Residual** | **68.546** |  |  |  |
| **STREM-1** | **Random** | **Time** | **5.934** |  |  |  |

The models assess the effect of sepsis diagnosis (pat_type), Time, and their interaction on log-transformed IFN-γ levels. Estimates (Estimate), standard errors (Std. Error), p-values, and 95% confidence intervals (Conf. low–Conf. high) are presented for fixed effects. Variance components are shown for random effects in Blue. Abbreviations: IL, interleukin; TNF-A, tumor necrosis factor alpha; TNFRSF1A, tumor necrosis factor receptor 1; IFNG, interferon gamma; CCL C-C motif chemokine ligand ; PCT, procalcitonin; STREM-11, soluble triggering receptor expressed on myeloid cells-1.

eTable 3**. Fixed and random effects estimates from mixed-effects models.**

| **Biomarker** | **Effect** | **Term** | **Beta Estimate** | **2.5% confidence limit** | **97.5% confidence limit** | **p value** |
| --- | --- | --- | --- | --- | --- | --- |
| IFNG | Fixed | (Intercept) | 1.091 | 1.003 | 1.179 | <2e-16 |
| IFNG | Fixed | SOFA score | -0.004 | -0.020 | 0.012 | 0.665 |
| IFNG | Fixed | Time | 0.001 | -0.001 | 0.003 | 0.582 |
| **IFNG** | **Random** | **(Intercept)** | **0.435** |  |  |  |
| **IFNG** | **Random** | **Residual** | **0.186** |  |  |  |
| **IFNG** | **Random** | **Time** | **0.005** |  |  |  |
| IL10 | Fixed | (Intercept) | 1.031 | 0.876 | 1.186 | < 2e-16 |
| IL10 | Fixed | SOFA score | 0.048 | 0.021 | 0.075 | 0.000818 |
| IL10 | Fixed | Time | -0.019 | -0.025 | -0.013 | 8.04e-11 |
| **IL10** | **Random** | **(Intercept)** | **0.783** |  |  |  |
| **IL10** | **Random** | **Residual** | **0.333** |  |  |  |
| **IL10** | **Random** | **Time** | **0.026** |  |  |  |
| IL17A | Fixed | (Intercept) | 0.675 | 0.612 | 0.738 | <2e-16 |
| IL17A | Fixed | SOFA score | 0.001 | -0.013 | 0.015 | 0.849 |
| IL17A | Fixed | Time | -0.001 | -0.003 | 0.001 | 0.398 |
| **IL17A** | **Random** | **(Intercept)** | **0.269** |  |  |  |
| **IL17A** | **Random** | **Residual** | **0.164** |  |  |  |
| **IL17A** | **Random** | **Time** | **0.006** |  |  |  |
| IL1B | Fixed | (Intercept) | 0.715 | 0.633 | 0.797 | <2e-16 |
| IL1B | Fixed | SOFA score | -0.012 | -0.030 | 0.006 | 0.179 |
| IL1B | Fixed | Time | -0.002 | -0.006 | 0.002 | 0.265 |
| **IL1B** | **Random** | **(Intercept)** | **0.335** |  |  |  |
| **IL1B** | **Random** | **Residual** | **0.263** |  |  |  |
| **IL1B** | **Random** | **Time** | **0.008** |  |  |  |
| IL1RN | Fixed | (Intercept) | 3.965 | 3.836 | 4.094 | < 2e-16 |
| IL1RN | Fixed | SOFA score | 0.044 | 0.017 | 0.071 | 0.00193 |
| IL1RN | Fixed | Time | -0.021 | -0.025 | -0.017 | 1.97e-15 |
| **IL1RN** | **Random** | **(Intercept)** | **0.572** |  |  |  |
| **IL1RN** | **Random** | **Residual** | **0.285** |  |  |  |
| **IL1RN** | **Random** | **Time** | **0.020** |  |  |  |
| IL6 | Fixed | (Intercept) | 2.249 | 2.086 | 2.412 | < 2e-16 |
| IL6 | Fixed | SOFA score | 0.048 | 0.009 | 0.087 | 0.015917 |
| IL6 | Fixed | Time | -0.015 | -0.023 | -0.007 | 0.000775 |
| IL6 | Fixed | Time:SOFA score | -0.003 | -0.005 | -0.001 | 0.027250 |
| **IL6** | **Random** | **(Intercept)** | **0.588** |  |  |  |
| **IL6** | **Random** | **Residual** | **0.442** |  |  |  |
| **IL6** | **Random** | **Time** | **0.014** |  |  |  |
| IL7 | Fixed | (Intercept) | 0.385 | 0.334 | 0.436 | <2e-16 |
| IL7 | Fixed | SOFA score | 0.006 | -0.004 | 0.016 | 0.2931 |
| IL7 | Fixed | Time | -0.002 | -0.004 | 0.000 | 0.0596 |
| **IL7** | **Random** | **(Intercept)** | **0.214** |  |  |  |
| **IL7** | **Random** | **Residual** | **0.118** |  |  |  |
| **IL7** | **Random** | **Time** | **0.007** |  |  |  |
| IL.8 | Fixed | (Intercept) | 2.200 | 2.075 | 2.325 | < 2e-16 |
| IL.8 | Fixed | SOFA score | 0.025 | -0.004 | 0.054 | 0.0842 |
| IL.8 | Fixed | Time | -0.012 | -0.018 | -0.006 | 9.96e-05 |
| **IL.8** | **Random** | **(Intercept)** | **0.435** |  |  |  |
| **IL.8** | **Random** | **Residual** | **0.415** |  |  |  |
| **IL.8** | **Random** | **Time** | **0.021** |  |  |  |
| CCL2 | Fixed | (Intercept) | 2.062 | 1.940 | 2.184 | < 2e-16 |
| CCL2 | Fixed | SOFA score | 0.046 | 0.021 | 0.071 | 0.000373 |
| CCL2 | Fixed | Time | -0.009 | -0.013 | -0.005 | 0.000130 |
| **CCL2** | **Random** | **(Intercept)** | **0.537** |  |  |  |
| **CCL2** | **Random** | **Residual** | **0.278** |  |  |  |
| **CCL2** | **Random** | **Time** | **0.016** |  |  |  |
| CCL7 | Fixed | (Intercept) | 0.878 | 0.813 | 0.943 | <2e-16 |
| CCL7 | Fixed | SOFA score | 0.000 | -0.014 | 0.014 | 0.949 |
| CCL7 | Fixed | Time | -0.001 | -0.003 | 0.001 | 0.549 |
| **CCL7** | **Random** | **(Intercept)** | **0.299** |  |  |  |
| **CCL7** | **Random** | **Residual** | **0.148** |  |  |  |
| **CCL7** | **Random** | **Time** | **0.006** |  |  |  |
| TNF.A | Fixed | (Intercept) | 1.035 | 0.917 | 1.153 | < 2e-16 |
| TNF.A | Fixed | SOFA score | 0.008 | -0.016 | 0.032 | 0.54104 |
| TNF.A | Fixed | Time | -0.006 | -0.010 | -0.002 | 0.00601 |
| **TNF.A** | **Random** | **(Intercept)** | **0.539** |  |  |  |
| **TNF.A** | **Random** | **Residual** | **0.262** |  |  |  |
| **TNF.A** | **Random** | **Time** | **0.016** |  |  |  |
| TNFRSF1A | Fixed | (Intercept) | 3.432 | 3.332 | 3.532 | < 2e-16 |
| TNFRSF1A | Fixed | SOFA score | 0.050 | 0.030 | 0.070 | 2.56e-06 |
| TNFRSF1A | Fixed | Time | -0.005 | -0.009 | -0.001 | 0.00661 |
| **TNFRSF1A** | **Random** | **(Intercept)** | **0.467** |  |  |  |
| **TNFRSF1A** | **Random** | **Residual** | **0.180** |  |  |  |
| **TNFRSF1A** | **Random** | **Time** | **0.016** |  |  |  |
| PCT | Fixed | (Intercept) | 1.345 | -5.392 | 8.082 | 0.695889 |
| PCT | Fixed | SOFA score | 2.630 | 1.321 | 3.939 | 0.000106 |
| PCT | Fixed | Time | 0.054 | -0.164 | 0.272 | 0.628450 |
| **PCT** | **Random** | **(Intercept)** | **31.213** |  |  |  |
| **PCT** | **Random** | **Residual** | **16.647** |  |  |  |
| **PCT** | **Random** | **Time** | **0.844** |  |  |  |
| STREM-1 | Fixed | (Intercept) | 297.498 | 28.935 | 566.061 | 0.0311 |
| STREM-1 | Fixed | SOFA score | -0.792 | -8.932 | 7.348 | 0.8490 |
| STREM-1 | Fixed | Time | 1.028 | -0.285 | 2.341 | 0.1263 |
| **STREM-1** | **Random** | **(Intercept)** | **1,633.077** |  |  |  |
| **STREM-1** | **Random** | **Residual** | **68.453** |  |  |  |
| **STREM-1** | **Random** | **Time** | **6.026** |  |  |  |

The models assess the effect of sofa scale, and Time, on log-transformed. IFN-γ levels. Variance components are shown for random effects in Blue. Abbreviations: CCL C-C motif chemokine ligand; IL, interleukin; TNF-A, tumor necrosis factor alpha; TNFRSF1A, tumor necrosis factor receptor 1; IFNG, interferon gamma; PCT, procalcitonin; STREM-1, soluble triggering receptor expressed on myeloid cells-1.

**eTable4. Results of joint modeling for longitudinal measures and Time to event.**

| Parameter | Hazard Ratio | Lower Credible Interval | Upper Credible Interval | P bayes |
| --- | --- | --- | --- | --- |
| edad | 1.073 | 1.025 | 1.128 | 0.002 |
| sofa_highest | 1.332 | 1.133 | 1.569 | 0.000 |
| IL10 | 1.082 | 0.999 | 1.197 | 0.053 |
| edad | 1.068 | 0.955 | 1.182 | 0.202 |
| sofa_highest | 1.415 | 1.025 | 1.856 | 0.042 |
| IL.8 | 0.655 | 0.582 | 0.728 | 0.000 |
| edad | 1.066 | 0.948 | 1.181 | 0.216 |
| sofa_highest | 1.444 | 1.042 | 1.892 | 0.035 |
| TNFRSF1A | 0.505 | 0.419 | 0.682 | 0.010 |
| edad | 1.085 | 1.032 | 1.145 | 0.002 |
| sofa_highest | 1.402 | 1.174 | 1.673 | 0.001 |
| PCT | 1.004 | 1.001 | 1.008 | 0.000 |
| edad | 1.073 | 1.002 | 1.144 | 0.047 |
| sofa_highest | 1.356 | 1.121 | 1.728 | 0.004 |
| IL1RN | 0.837 | 0.513 | 1.025 | 0.153 |
| edad | 1.079 | 1.024 | 1.147 | 0.006 |
| sofa_highest | 1.385 | 1.156 | 1.684 | 0.002 |
| IL6 | 0.799 | 0.462 | 1.342 | 0.434 |
| edad | 1.081 | 1.027 | 1.143 | 0.003 |
| sofa_highest | 1.337 | 1.120 | 1.600 | 0.002 |
| CCL2 | 1.223 | 0.972 | 1.628 | 0.081 |
| edad | 1.076 | 1.026 | 1.130 | 0.001 |
| sofa_highest | 1.281 | 1.095 | 1.495 | 0.001 |
| STREM-1 | 1.000 | 0.999 | 1.000 | 0.812 |

Results are obtained by joint model analysis, corrected for age, and the highest SOFA at the day of sampling. Joint model analysis was used to evaluate the longitudinal changes in plasma biomarker concentrations and the association between changes in biomarker concentration and outcomes. Joint modelling analyses two types of data: repeated measurements over Time and Time-to-event data (survival data). P_Bayes_ indicates the posterior probability that the parameter differs from zero, as estimated from the joint Bayesian models. Credible intervals represent the ranges containing the true parameter value with 95% posterior probability. Abbreviations: IL, interleukin; TNFRSF1A, tumor necrosis factor receptor 1; MCP, monocyte chemoattractant protein; PCT, procalcitonin; STREM-1, soluble triggering receptor expressed on myeloid cells-1.

**eFig 6 ROC curves and AUC of Joint modeling evaluating, IL-8, TNF-RI and PCT**

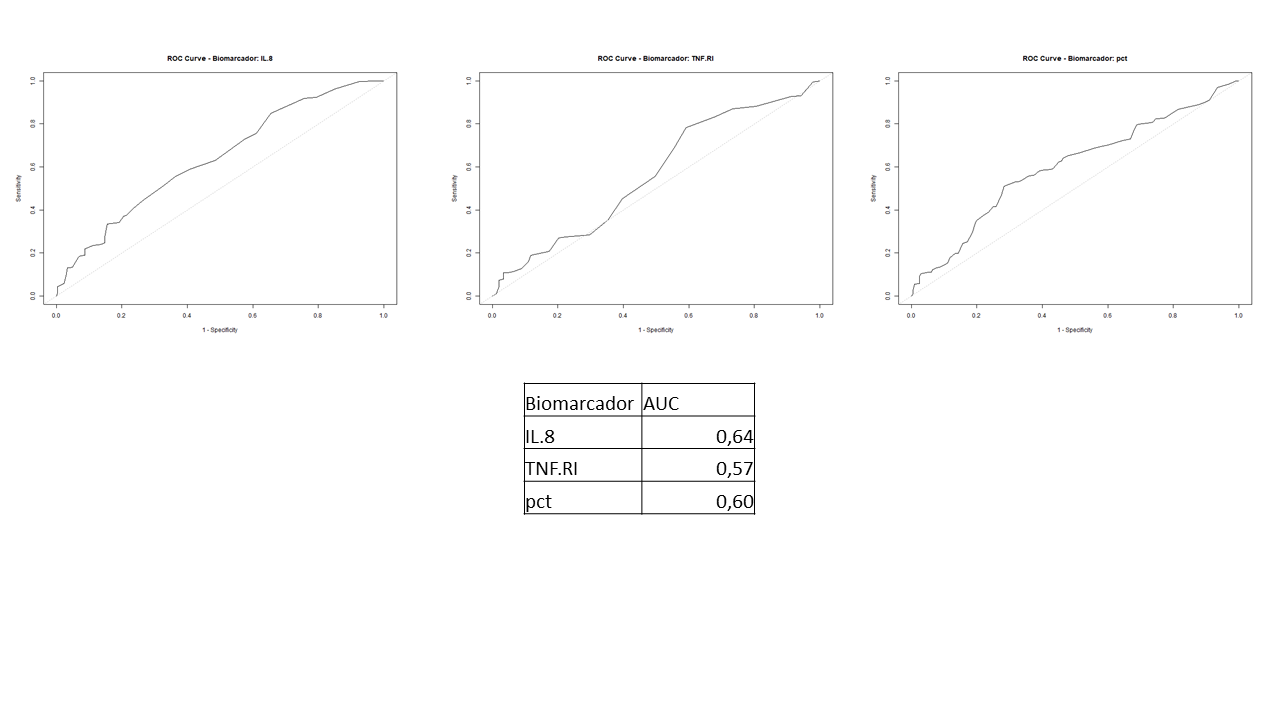

ROC curves evaluating the ability to predict the event at 1024 hours based on data from 24 hours.

**eFig 7 Calibration Plot and Brier scores of Joint modeling evaluating IL8, TNF-RI and PCT.**

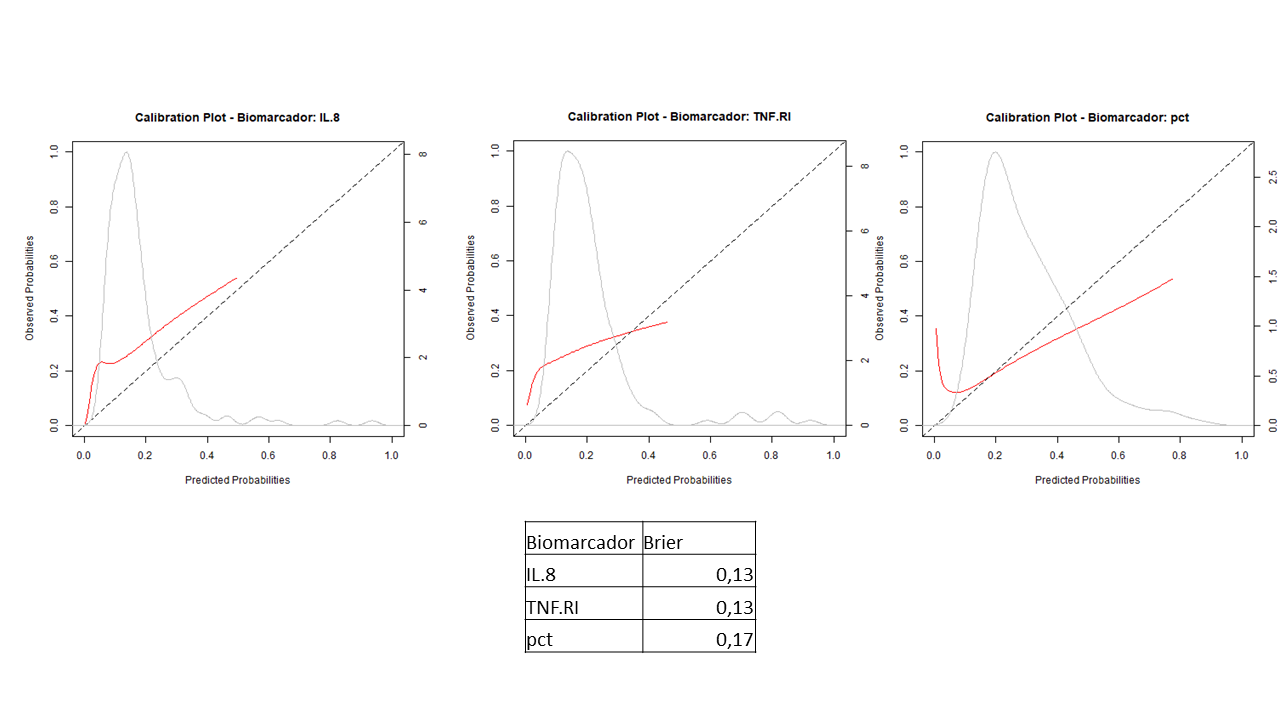

Calibration curves and Brier scores evaluating the predictive performance of the joint models using longitudinal data up to 24 hs with horizon time at 524 hours after ED admission. The plots depict the agreement between predicted and observed event probabilities across risk deciles, while Brier scores quantify overall accuracy over the respective prediction horizons.
